# Slight reduction in SARS-CoV-2 exposure viral load due to masking results in a significant reduction in transmission with widespread implementation

**DOI:** 10.1101/2020.09.13.20193508

**Authors:** Ashish Goyal, Daniel B. Reeves, E. Fabian Cardozo-Ojeda, Bryan T. Mayer, Joshua T. Schiffer

## Abstract

Masks are a vital tool for limiting SARS-CoV-2 spread in the population. Here we utilize a mathematical model to assess the impact of masking on transmission within individual transmission pairs and at the population level. Our model quantitatively links mask efficacy to reductions in viral load and subsequent transmission risk. Our results reinforce that the use of masks by both a potential transmitter and exposed person substantially reduces the probability of successful transmission, even if masks only lower exposure viral load by ∼50%. Slight increases in mask adherence and/or efficacy above current levels would reduce the effective reproductive number (R_e_) substantially below 1, particularly if implemented comprehensively in potential super-spreader environments. Our model predicts that moderately efficacious masks will lower exposure viral load 10-fold among people who get infected despite masking, potentially limiting infection severity. Because peak viral load tends to occur pre-symptomatically, we also identify that antiviral therapy targeting symptomatic individuals is unlikely to impact transmission risk. Instead, antiviral therapy would only lower R_e_ if dosed as post-exposure prophylaxis and if given to ∼50% of newly infected people within 3 days of an exposure. These results highlight the primacy of masking relative to other biomedical interventions under consideration for limiting the extent of the COVID-19 pandemic prior to widespread implementation of a vaccine.

## Introduction

Masks are a barrier method to prevent the spread of respiratory viral infections. A mask essentially serves as a filter that prevents passage of some portion of viruses from the airway of the transmitter to the airway of exposed contacts. Mask efficacy is therefore mediated by a reduction in exposure viral load (*1, 2*). If a potential transmitter as well as an exposed contact are masked, then this filtering process occurs twice potentially amplifying protection.

Mask efficacy is inferior to that of other, less permeable barrier methods to prevent infections such as condoms (*1, 3, 4*). The most commonly used cloth and hospital masks do not provide a perfect facial seal and mask fabric does not block emission or inhalation of all aerosolized viral particles (*5, 6*). N95 masks may bypass these shortcomings but are in short supply and difficult to wear for long periods of time (*7, 8*). As a result of these imperfections, recommendations for mask use have varied over the course of the COVID-19 pandemic. Nevertheless, widespread mask use is recognized as a critical component of any viable public health strategy against COVID-19 (*9-11*). Recent models demonstrate that slight increases in mask utilization could be the single most important factor that prevents exponential growth in incident cases (*12-14*).

Quantifying mask efficacy in real-world settings remains challenging. Elegant experimental work demonstrated the efficacy of masks in animal models (*15*). Many studies have been performed in hospital settings where mask compliance is uniform and other complementary infection prevention methods are more commonly employed than in other public gathering or work locations (*16, 17*). To the best of our knowledge, no study has captured the impact of masking on the likelihood of super-spreader events, with specific consideration of intermittent compliance.

Here we develop a mathematical model capturing viral load-mediated effects of mask use on transmission probability within transmission pairs and at the population level. We use this approach to estimate the efficacy of masks in real world settings, and to characterize effects on super-spreader events as well as exposure viral loads of those who get infected despite masking. Finally, we compare the preventative impact of masking to the use of antiviral therapies given early during symptomatic infection, or when used as post-exposure prophylaxis (PEP).

## Results

### Baseline mathematical model of SARS-CoV-2 viral load dependent transmission

We employed a previously developed mathematical model which links viral load shedding at the individual level with population level epidemic spread (*18, 19*), to determine the impact of masks on epidemics. Briefly, the model is built upon the assumptions that each transmitter has a specific number of exposure contacts per day and that each exposure contact has a certain probability of successful transmission based on the viral load of the transmitter. This probability is based on a transmission dose (TD) response curve. We fit the model to frequency histograms describing heterogeneity of individual R0, or the number of secondary infections attributed to each infected person (*20-24*). The individual R0 distribution for SARS-CoV-2 transmission is highly over-dispersed, meaning that most infected people do not spread infection while a minority infect a large number of people. We also fit the model to distributions of individual serial intervals, the time from the onset of symptoms in the transmitter to symptom onset in the secondarily infected person (*25*). The overall qualitative conclusions of this model were that the period of contagiousness for SARS-CoV-2 is quite short, typically less than a day, and that super-spreader events are largely attributable to high variability in the number of exposure contacts per day among infected people.

### Predicted impact of transmitter or exposure contact masking on transmission probability within transmission pairs

We added masking to this model by assuming that a mask decreases the exposure viral load in a transmission pair by a value that we refer to as the combination mask efficacy (*ε*_*C*_). This efficacy represents the proportion of viruses filtered by masks worn by both the transmitter and exposed person. If the transmitter is wearing mask with efficacy *ε*_*T*_ and the exposed person is wearing a mask with efficacy *ε*_*E*_, then the exposure viral load *V*_*E*_ can be related to the transmitter viral load *V*_*T*_ by: *V*_*E*_ = *V*_*T*_ (*1-ε*_*T*_)(*1-ε*_*E*_). The combination mask efficacy is then *ε*_*C*_=*1-*(*1-ε*_*T*_)(*1-ε*_*E*_), which takes on a value of zero when both parties are not wearing a mask or wearing masks that are totally ineffective **(Fig 1)**.

**Figure 1.**
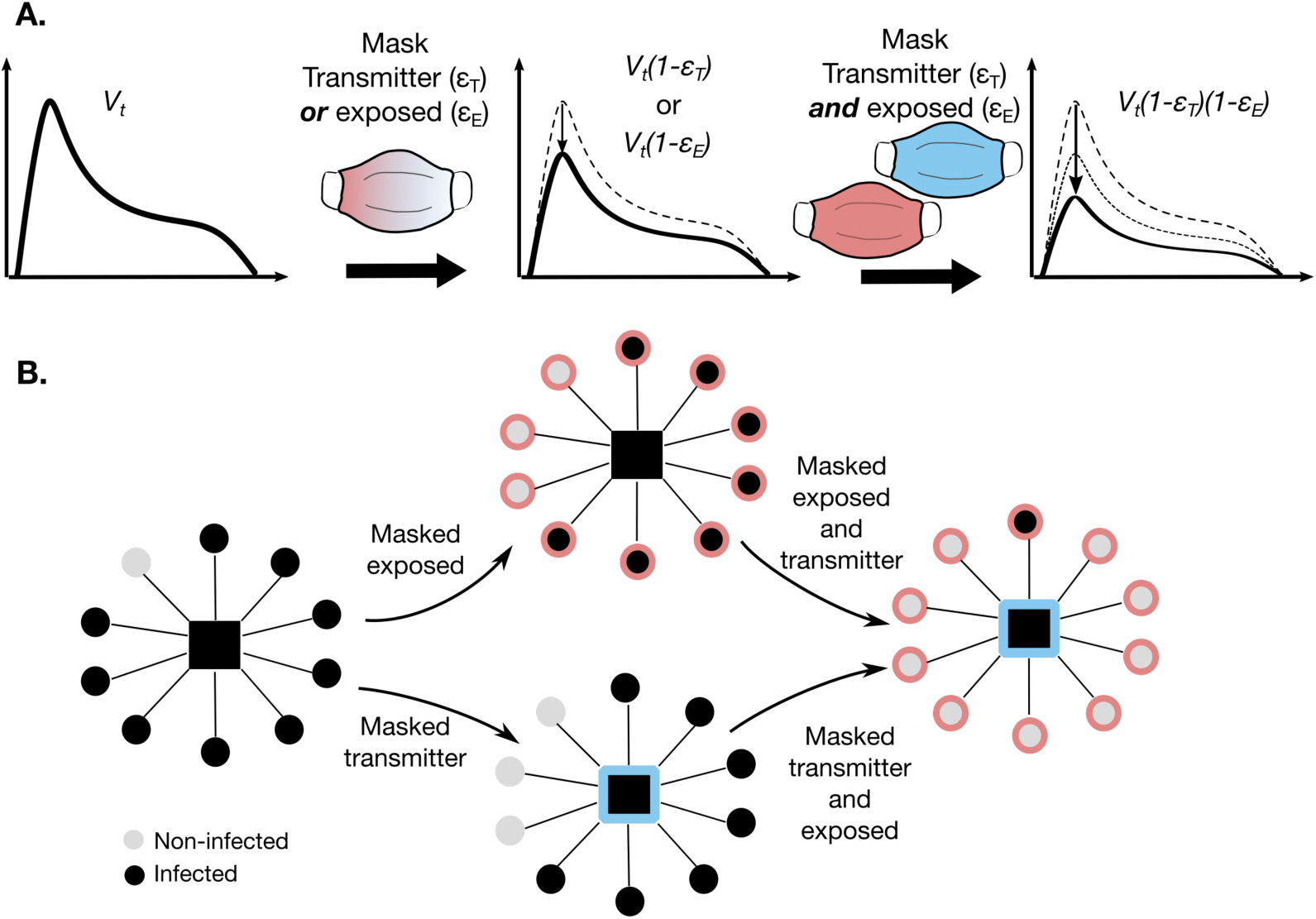
Mathematical model of mask impact on SARS-CoV-2 exposure viral load. **a**. The viral load emitted by a potential transmitter (V_t_) can be filtered, resulting in lower exposure viral loads due to a single mask worn by a transmitter or exposed individual with efficacy eT or eE respectively. Dual masking lowers exposure viral load further by filtering virus twice. **b**. Dual masking may prevent super-spreader events to a greater extent than a masked transmitter or masked exposed individual.

As in our prior model, the exposure viral load impacts contagiousness, which is the probability that virus is passaged to the exposed person’s airway, as well as infectiousness, the probability of cellular infection given the presence of virus in the airway. Each of these properties is associated with a dose response curve (contagiousness dose (CD) response curve and infectiousness dose (ID) response curve), the product of which is the TD response curve.

In this context, we first establish a baseline probability of transmission given no use of masks on both sides of a potential transmission pair **(Fig 2a)**. The absolute & relative reductions in transmission probability with more effective masks vary as *V*_*T*_ increases. At lower viral loads (<10^7.5^ viral RNA copies), a moderate to highly effective mask worn by either a transmitter (0.9>*ε*_*T*_≥0.5) or an exposed contact (0.9>*ε*_*E*_≥0.5) is sufficient to partially lower the absolute probability of transmission **(Fig 2b-c, Sup fig 1a)**. The relative reduction in transmission probability increases linearly with increasing mask efficacy at 10^7^ viral RNA copies with an increasingly concave, curvilinear relationship at higher viral loads **(Sup fig 1c)**.

**Figure 2.**
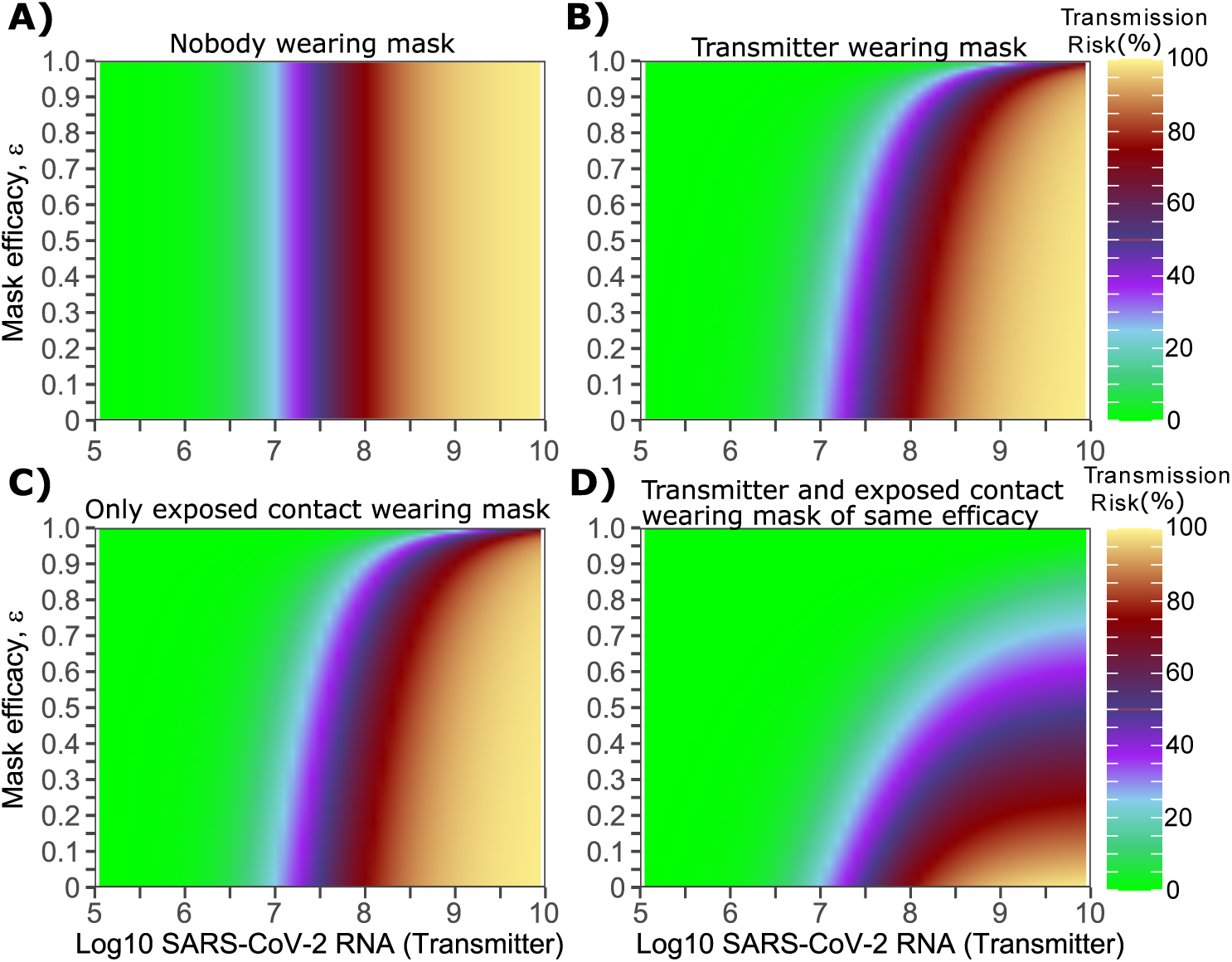
Impact of masking of the transmitter alone, the exposed contact alone or both members of the transmission pair, on transmission risk given a single exposure contact. **a-d**. Each panel is based on simulations of 1000 transmission pairs. **a**. No masking, **b**. Transmitter is masked, **c**. Exposed contact is masked, **d**. Both members are masked.

At higher transmitter viral load (10^7.5^–10^9^ viral RNA copies), a moderate to highly effective mask worn by either a transmitter (0.9>*ε*_*T*_≥0.5) or an exposed contact (0.9>*ε*_*E*_≥0.5) insignificantly lowers the absolute probability of transmission **(Fig 2b-c, Sup fig 1a)**. At high viral loads, the relative reduction in transmission probability increases dramatically with extremely effective masks of efficacy ≥0.9, when the mask is worn by either a transmitter or an exposed contact **(Sup fig 1c)**.

### Predicted impact of dual masking on transmission probability within transmission pairs

If both the transmitter and exposed person wear masks (dual masking), then lower mask efficacies are sufficient to significantly lower transmission risk at a wider range of exposure viral loads **(Fig 2d)**. At viral loads <10^8^ viral RNA copies, masks worn by both transmitter and exposed contact of more than moderate efficacy (*ε*_*T*_≥0.5, *ε*_*E*_≥0.5 resulting in *ε*_*C*_≥0.75), is sufficient to partially lower the absolute probability of transmission **(Fig 2d, Fig 3, Sup fig 1b)**. The relative reduction in transmission probability according to mask efficacy increases more rapidly with dual **(Sup fig 1d)** compared to single **(Sup fig 1c)** masking. If both transmitter and exposed contacts wear masks with *ε*_*T*_= 0.9 and *ε*_*E*_ = 0.9 (*ε*_*C*_=0.99), then transmission probability is reduced to <5% for viral loads <10^8.5^ viral RNA copies and to ∼20% for transmitter viral load of 10^9^ viral RNA copies **(Fig 2d, 3d & Sup fig 1c)**.

**Figure 3.**
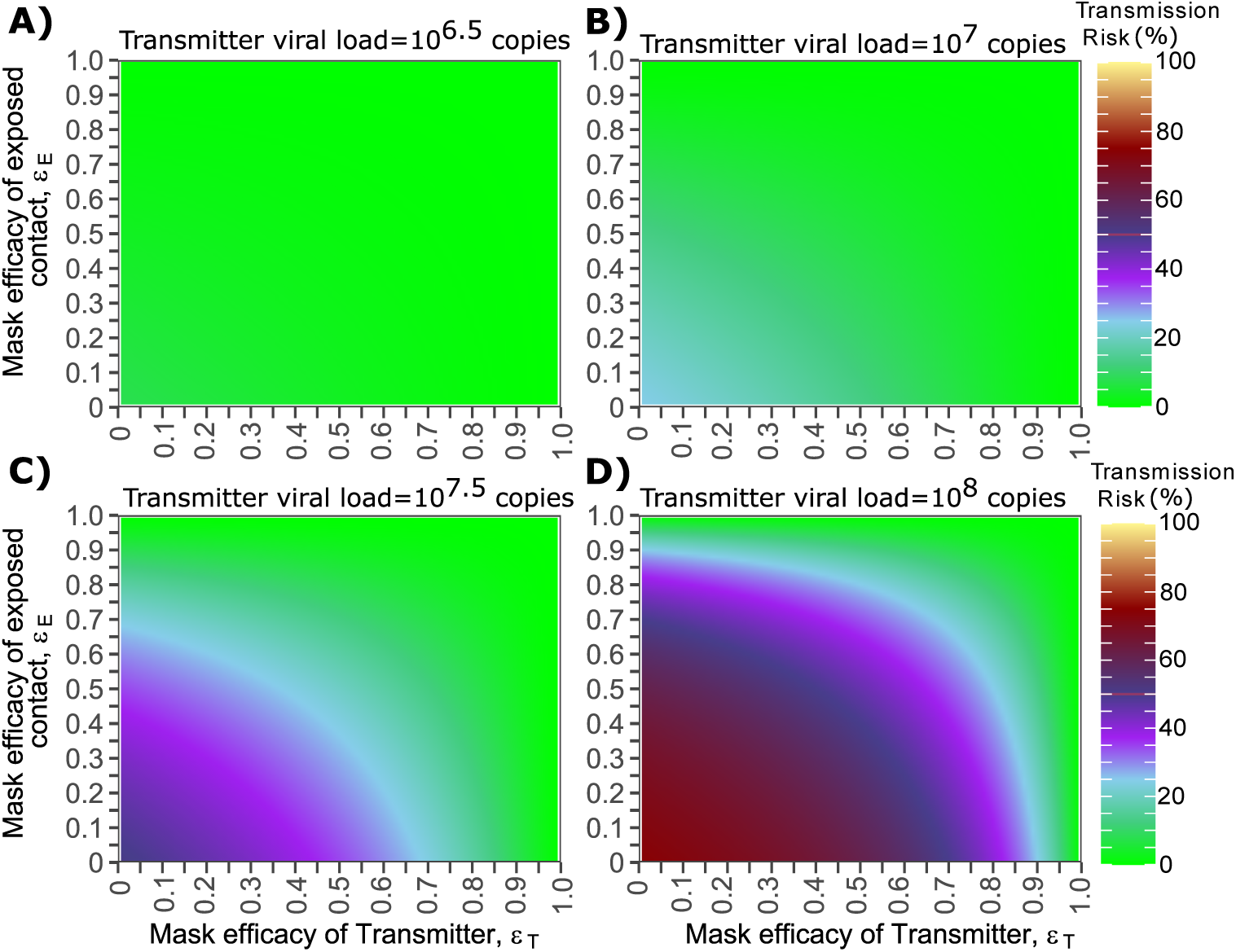
Impact of masking of both members of a transmission pair on transmission risk according to transmitter viral load. **a-d**. Each panel is based on simulations of 1000 transmission pairs. **a**. Low viral load, **b**. Moderate viral load, **c**. High viral load, **d**. Extremely high viral load.

### Predicted impact of transmitter and exposure contact masking on effective reproduction number (R_e_) at different levels of implementation

We next explore the impact of general masking adherence rates on population level metrics of infection by simulating 3000 potential transmitters assuming heterogeneity in viral load trajectories and exposure contact networks among individuals. Reduction in the effective reproductive number (R_e_) depends on both the mask efficacy levels (*ε*_*T*_ and *ε*_*E*_) and the level of adherence to masking **(Fig 4)**. If we assume 25% of people wear masks 25% of the time, then most variability in results occur due to the stochastic nature of the model **(Fig 4a)**. If we assume that 50% of people wear masks 50% of the time, then the use of masks with high efficacy of ∼0.9 results in a drop of R_e_ from ∼1.8 to ∼1.0 **(Fig 4b)**. With 75% of people wearing masks 75% of the time, a mask efficacy of ∼0.5 allows for a reduction of R_e_ from ∼1.8 to ∼1.0 **(Fig 4c)**. With 100% of people wearing masks 100% of the time, then a mask efficacy of ∼0.3 is sufficient to achieve R_e_ ∼1.0, and efficacy of 0.5 in both transmitter and exposed contacts lowers R_e_ to less than 0.6 **(Fig 4d)**.

**Figure 4.**
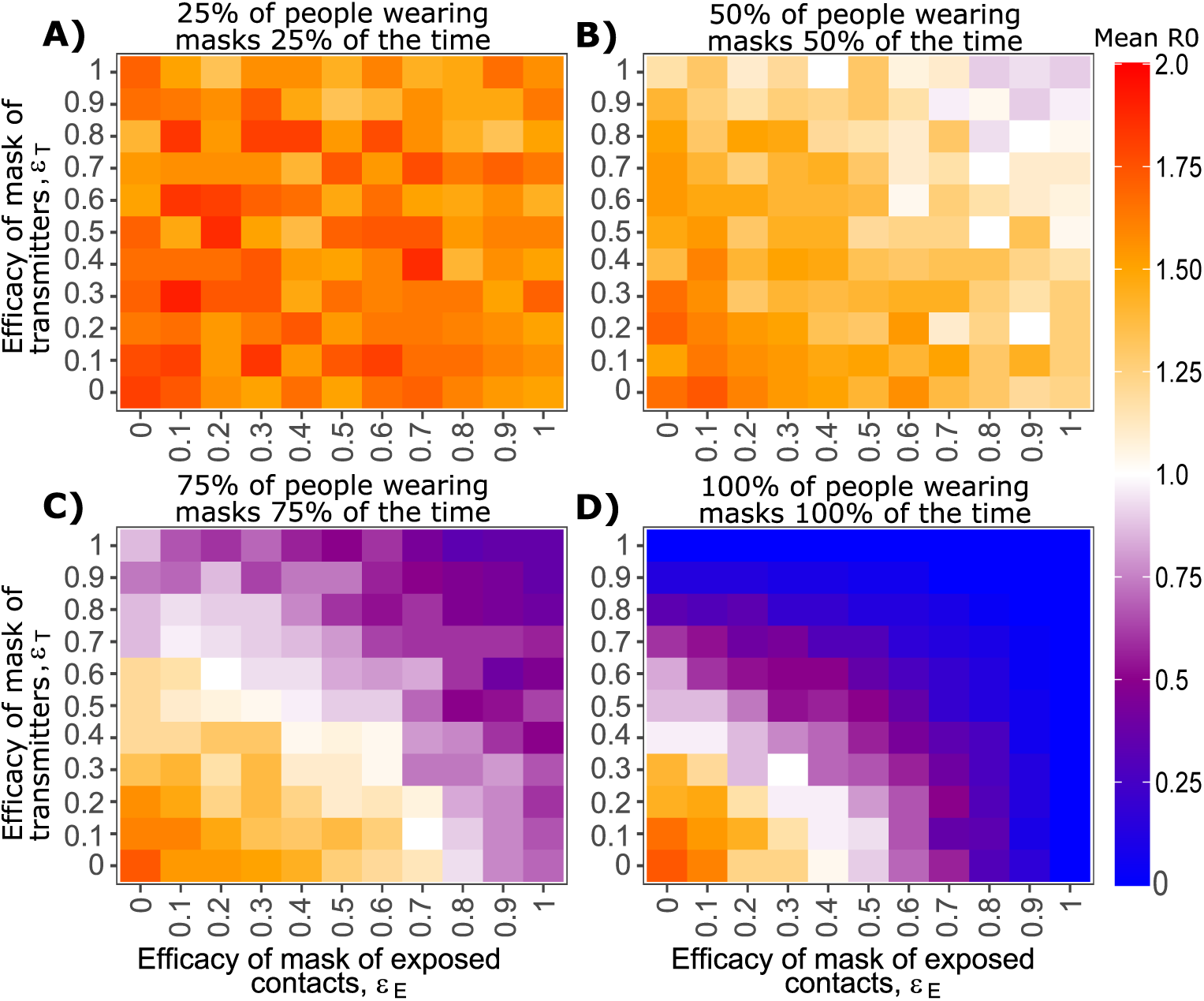
Effect of mask utilization and efficacy on mean SARS-CoV-2 R0. Each heat map is based on simulations of 3000 transmitters with varying daily exposure contacts. **a**. Low mask utilization, **b**. Moderate mask utilization, **c**. High mask utilization, **d**. Extremely high mask utilization.

Current estimates of daily mask use over the last 2 months vary between states in the United States between 40 and 60% (http://www.healthdata.org/acting-data/maps-mask-use), and R_e_ has varied between 0.8 and 1.2 (https://covid19-projections.com/us). These results suggest that panel **Fig 4c** is likely the closest to current U.S. epidemic conditions and that ε_T_ and ε_E_ likely fall roughly between 0.4 and 0.6 in a real-world setting, if mask efficacy is equivalent between transmitter and exposed contacts. While a combined efficacy *ε*_*C*_ of 0.65-0.85 can be roughly estimated from the model, the possibility of superior efficacy of masks in transmitters versus exposed, or vice versa, cannot be excluded.

### Proportions of infections attributable to masked and unmasked transmission pairs

We next project the proportion of transmission events attributable to different masking profiles among transmission pairs assuming equally effective mask (*ε*) used by both transmitters and exposed contacts. In circumstances with low mask utilization (25% of people wearing masks 25% of the time), nearly all transmissions occur from an unmasked person to an unmasked person **(Fig 5a)**. A similar trend is noted for moderate mask utilization (50%), particularly as mask efficacy increases **(Fig 5b)**.

**Figure 5.**
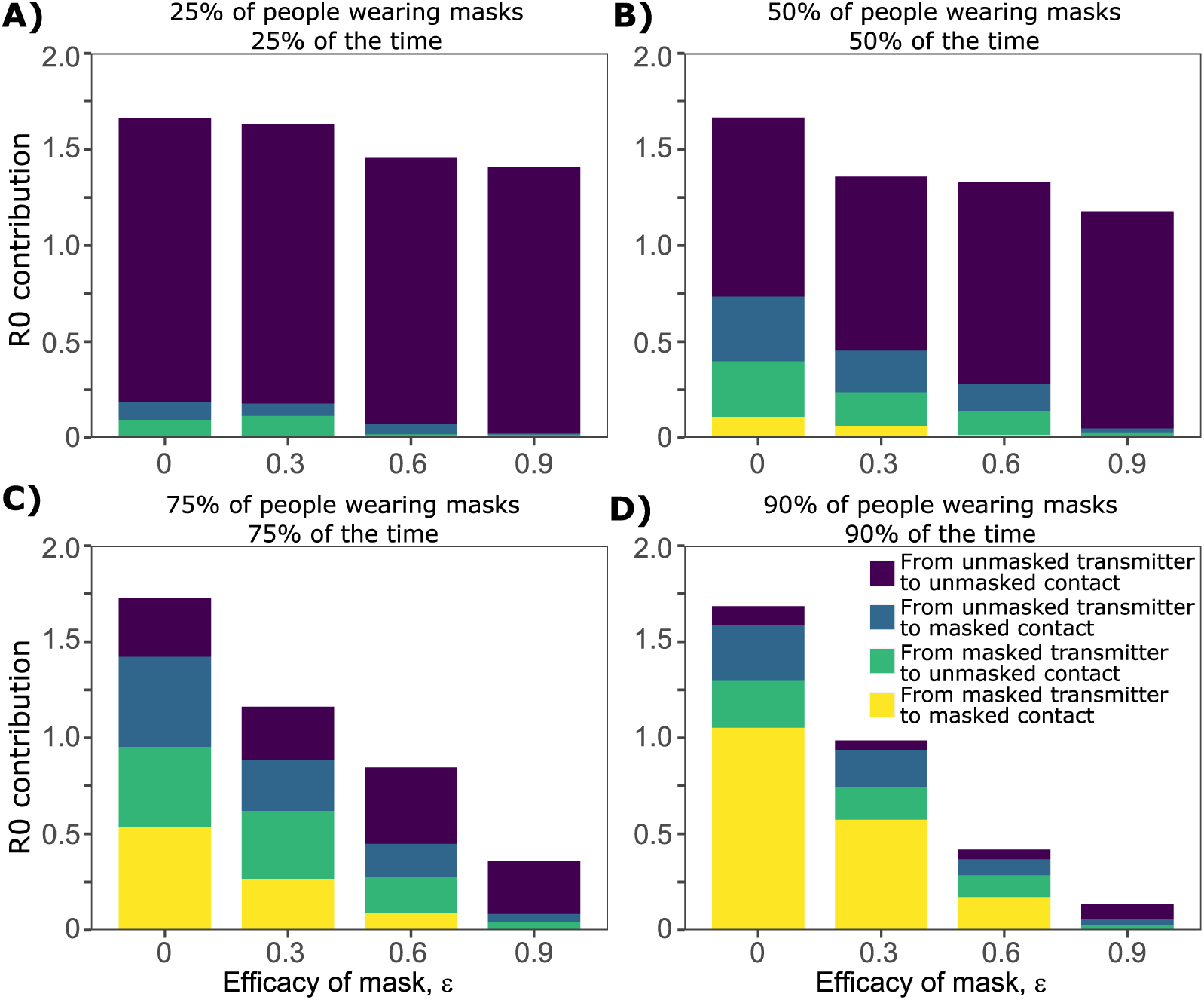
Effect of mask utilization and efficacy on proportion of masked transmissions contributing to total R0. Each histogram bar in each panel is based on simulations of 3000 transmitters with varying daily exposure contacts. **a**. Low mask utilization, **b**. Moderate mask utilization, **c**. High mask utilization, **d**. Extremely high mask utilization.

For high (75%) and extremely high (90%) mask utilization scenarios, if mask efficacy is moderate (*ε*∼0.6) as is currently believed, then a higher proportion of transmissions occur to or from a person wearing a mask, despite the fact that the total number of transmissions dramatically decreases **(Fig 5c, d)**.

We project that at current epidemic conditions in the United States, including ∼50% mask use (similar to 75% of people wearing masks 75% of the time) and mask efficacy of 0.3-0.6, that fewer than half of ongoing transmissions occur within unmasked pairs and that transmission between masked transmitters or masked exposed contacts likely contributes significantly to R_e_ **(Fig 5c)**.

### Predicted impact of transmitter and exposure contact masking on super-spreader events

We next identified that increased mask compliance and efficacy dramatically decreases the proportion of infected people who successfully transmit to another person **(Fig 6a-d)**. If 50% of people were to wear 50% effective masks half of the time, then the likelihood of an individual transmitting decreases from 30% to 20% **(Fig 6b)**. If mask compliance is increased to 75% of people 75% of the time, then the likelihood of a person transmitting decreases to ∼15% **(Fig 6c)**.

**Figure 6.**
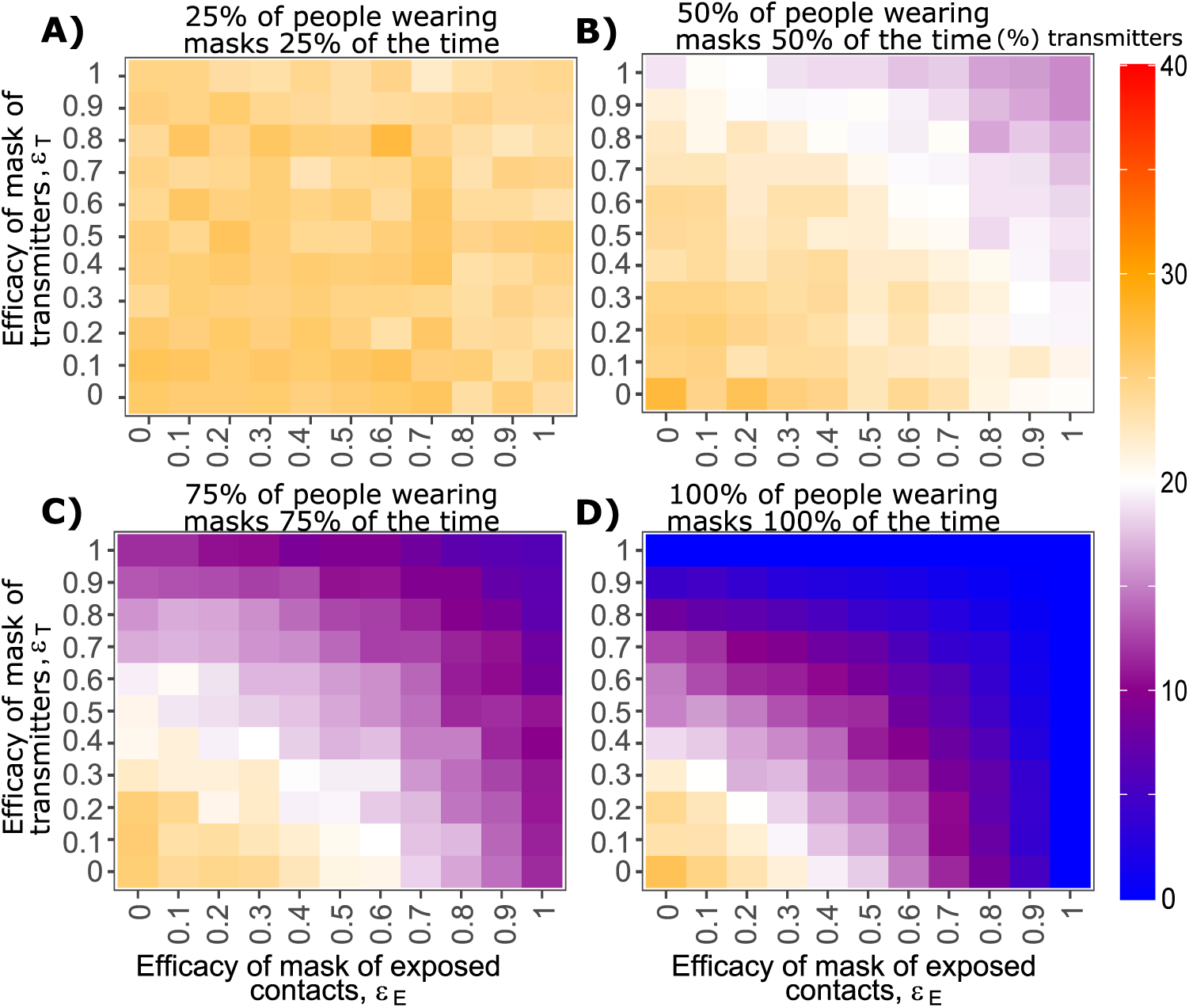
Effect of mask utilization and efficacy on percent of infected people who transmit to others. Each heat map is based on simulations of 3000 transmitters with varying daily exposure contacts. **a**. Low mask utilization, **b**. Moderate mask utilization, **c**. High mask utilization, **d**. Extremely high mask utilization.

When masking is applied homogeneously across the population, the proportion of infectors (transmitters who infect at least one person) who pass the infection to 5 or more people decreases, as mask utilization and efficacy increase **(Sup fig 2a-d)**. Increased mask utilization and increased mask efficacy leads to an even reduction of all types of transmission events, including transmissions to small numbers (1-3) of people, or super-spreader events to >5, >10, >20 or >50 people **(Fig 7a-d)**. Improvements in mask efficacy have a larger impact as utilization of mask use increases, including against super-spreader events **(Fig 7c, d)**. Under all simulations, super-spreader events with transmission to >5 people persist and make a nearly equivalent contribution proportionally to overall R_e_, though their absolute impact is considerably lessened with higher mask compliance and efficacy.

**Figure 7.**
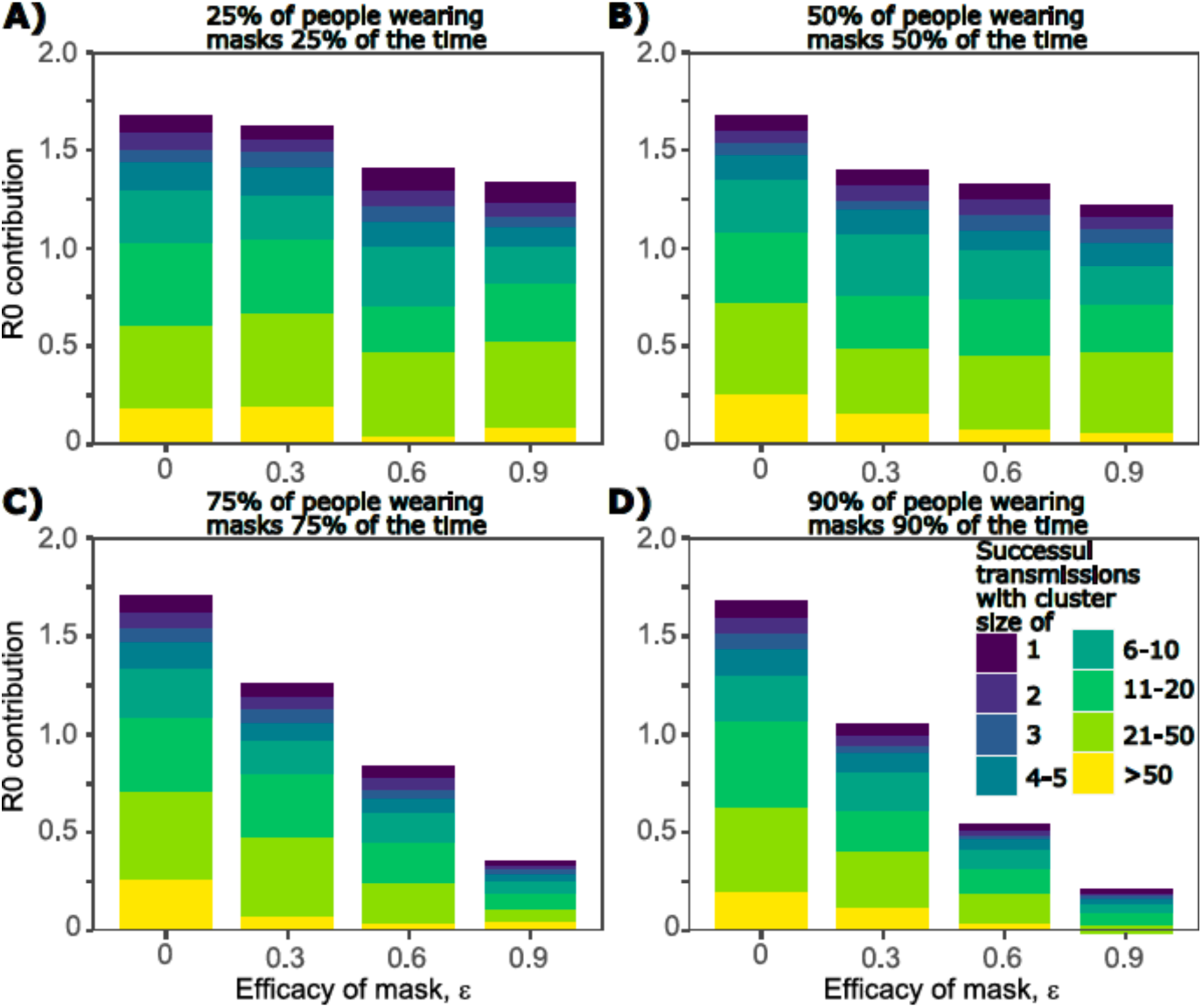
Effect of mask utilization and efficacy on proportion of super-spreader transmissions. Each histogram bar in each panel is based on simulations of 3000 transmitters with varying daily exposure contacts. **a**. Low mask utilization, **b**. Moderate mask utilization, **c**. High mask utilization, **d**. Extremely high mask utilization.

Our results suggest that with current levels of masking in the United States **(Fig 7c**, *ε*= 0.3-0.6), most of the contribution to R_e_ still comes from super-spreader events involving >5 secondary infections. We therefore simulated masking applied to 100% of people with >10 exposure contacts per day **(Sup fig 3a-d)** and found that even modest uptake of moderately effective masks (∼0.5) in the remainder of the population appeared to be sufficient to maintain R_e_ <1.

### Predicted impact of transmitter and exposure contact masking on viral inoculum at time of infection

Another theoretical benefit of masks is reduction in exposure viral load which in animal models of SARS-CoV-1 and MERS, leads to less severe infection (*26-28*). Simulating under the assumption that 75% of people wear masks 75% of the time (i.e., a situation representing current levels of masking in the United States), we identified that viral load of a transmitter required to generate secondary infections increases slightly with higher implementation of more efficacious masks, particularly with dual masking of transmitters and exposed contacts **(Fig 8a)**. Viral load exposure at the time of a successful transmission decreased according to efficacy of mask, particularly if both transmitters and exposed contacts are masked **(Fig 8b)**. With dual masking in place with efficacies of 0.6, exposure viral load decreased by ∼1 log **(Fig 8b)**. With dual masking in place with efficacies of 0.9, exposure viral load decreased by ∼2 logs **(Fig 8b)**.

**Figure 8.**
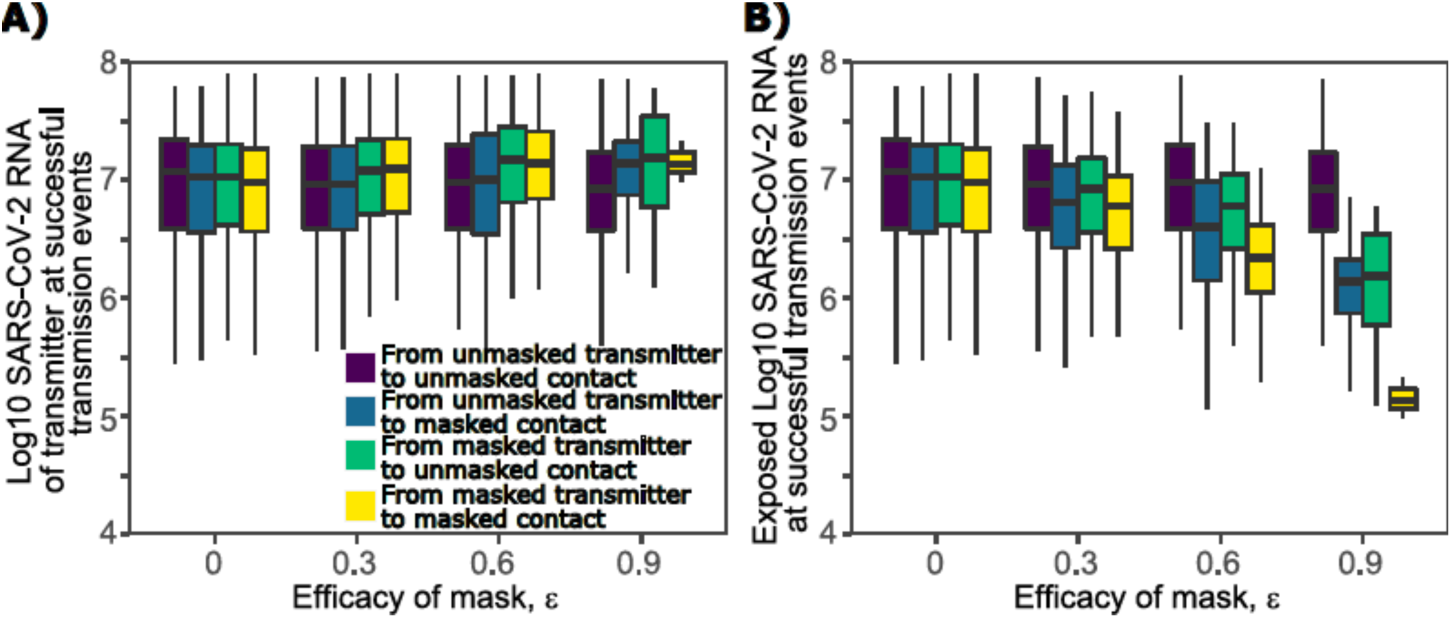
Effect of mask efficacy on exposure viral load during successful transmissions. Each boxplot in each panel is based on simulations of 3000 transmitters with varying daily exposure contacts. Boxplots are median and interquartile range (IQR) and lines are 1.5 the IQR. **a**. Viral load of transmitters, **b**. Viral load exposure in secondarily infected persons.

### Predicted impact of antiviral therapy during early symptomatic infection on R_e_

Treatment as prevention is a highly effective means for reducing person-to-person transmission of HIV (*29, 30*). Our models predict that initiation of potent antiviral therapy early after symptom onset is likely to have therapeutic benefit (*18*). We therefore tested whether early symptomatic therapy which rapidly eliminates shedding might also decrease secondary transmissions. Owing to the fact that symptomatic therapy would almost invariably occur after peak viral shedding **(Sup Fig 4a)**, our simulations suggest that even widespread implementation of early symptomatic therapy would not lower R_e_ **(Fig 9a)**, percentage of infected people who transmit to at least one other person **(Fig 9c)** or percentage of infectors who transmit to at least 5 other people **(Fig 9e)**.

**Figure 9.**
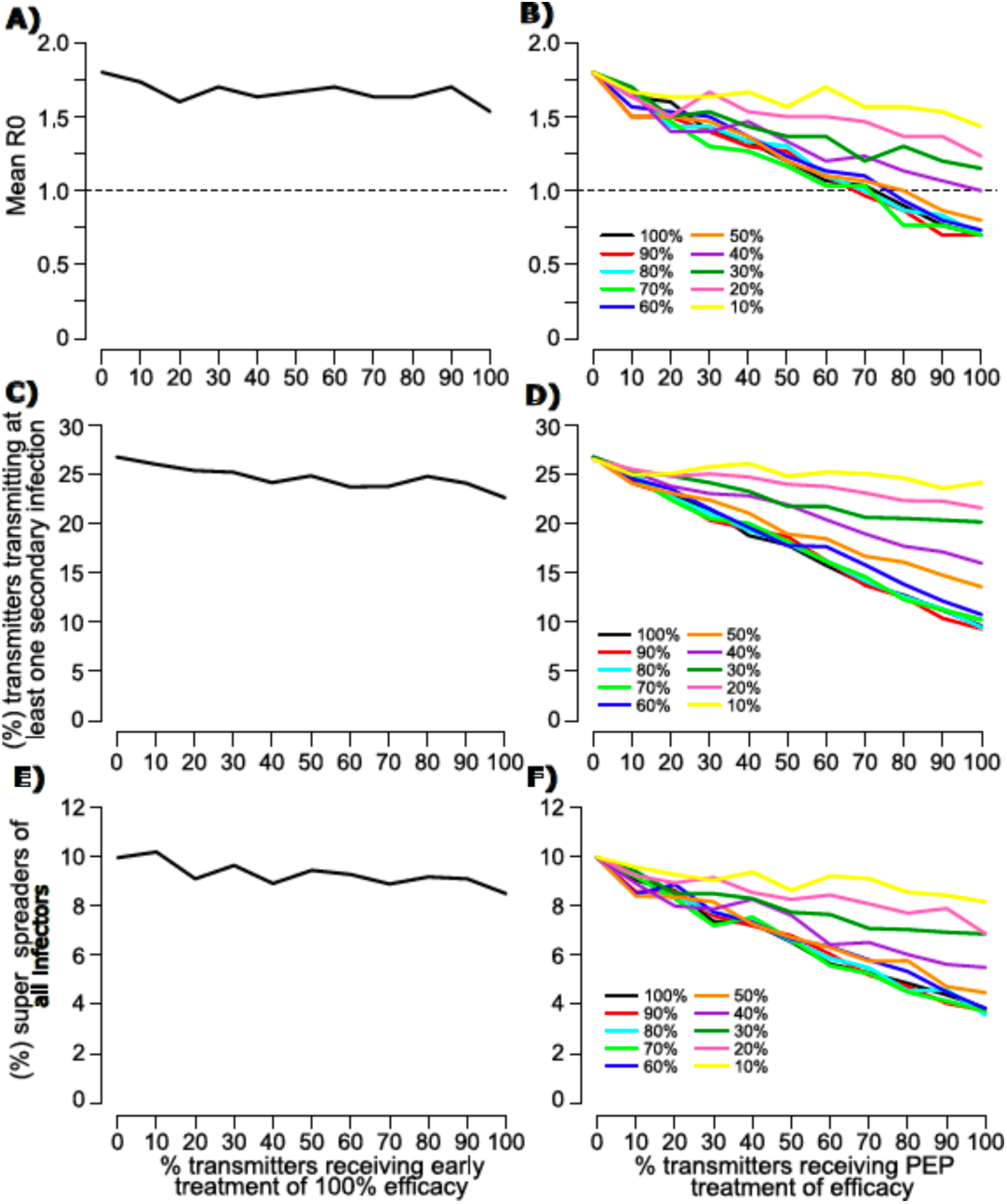
Projected impact of antiviral therapy on SARS-CoV-2 R0. Each line in each panel is based on simulations of 3000 transmitters with varying daily exposure contacts. Panels a, c and e are for early symptomatic therapy. Each simulation assumes 100% efficacy. Panels b, d and f are for post-exposure prophylaxis given during pre-symptomatic infection. Colored lines assume different antiviral efficacies (inset). **a, b**. Projected R0 given different amounts of uptake in the population. **c, d**. Projected percent of transmitters who infect at least one person given different amounts of uptake in the population. **e, f**. Projected percent of infectors who infect at least five people given different amounts of uptake in the population.

### Predicted impact of PEP on R_e_

PEP is also a potential method for lowering SARS-CoV-2 transmissions. Because PEP is given in the pre-symptomatic phase and would usually fall before or near peak viral shedding **(Sup Fig 4b)**, our simulations suggest an inverse linear relationship between uptake of PEP and R_e_ **(Fig 9b)**, percentage of infected people who transmit to at least one other person **(Fig 9d)** or percentage of infectors who transmit to at least 5 other people **(Fig 9f)**. To achieve R_e_ <1 would require PEP efficacy of 50% and ∼75% uptake in the population, which would in turn require 75% of SARS-CoV-2 cases to be contact traced. Increases in PEP efficacy beyond 0.5 would provide minimal to no enhancement of this benefit **(Fig 9b)**.

We further determine that the timing of PEP is critical. If PEP is initiated within 2 days of exposure, then the percent of people receiving effective treatment is highly predictive of R_e_ **(Fig 10)**. However, from day 3 onwards, the impact of effective PEP diminishes **(Fig 10)**.

**Figure 10.**
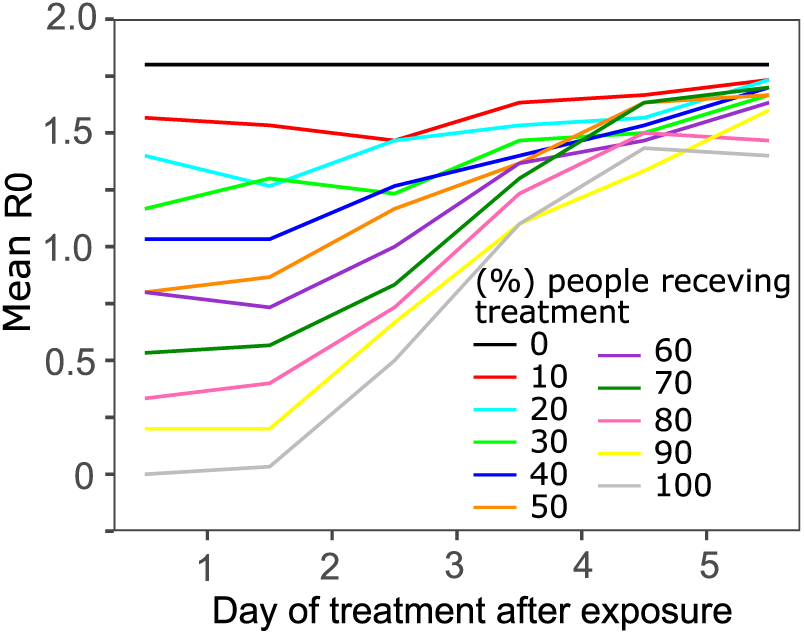
Projected impact of post-exposure prophylaxis on SARS-CoV-2 according to the day of implementation. Each line in is based on simulations of 3000 transmitters with varying daily exposure contacts and are colored by percent of transmitters receiving therapy.

Overall, these results highlight the fact that masking is likely to have more of an impact on R_e_ than any form of licensed antiviral therapy that emerges during the course of the pandemic.

## Discussion

Relative to other barrier methods for preventing the spread of infectious diseases such as condoms, masks are imperfect (*31*). Surgical and cloth masks, which are now used commonly by members of the public, do not completely eliminate droplet and airborne emission of viral particles by a transmitter (*1*). Nor do they prevent viral exposure to airway cells among exposed contacts. Their effectiveness in real world settings is further limited by intermittent compliance and improper masking technique. As a result, masks only prevent a proportion of person-to-person transmissions.

Nevertheless, our results demonstrate that in the absence of a licensed vaccine, based on moderate efficacy, low cost, high availability, and ease of use, masks are the most effective currently available biomedical intervention. If implemented widely and strategically, on top of baseline levels of physical distancing, masking would be sufficient to suppress ongoing spread of SARS-CoV-2 until widespread deployment of a vaccine is possible. More specifically, our model suggests that increased masking would lower the effective reproduction number (R_e_), lower the percentage of infected people who transmit the virus, decrease the total number of super-spreader events, and lower the exposure viral loads among infected people, possibly leading to less severe infections overall (*26*).

Importantly, there appears to be a critical threshold of compliance. We predict massive additional benefits accrued with an increase in masking compliance from 75% of people masking 75% of the time to 90% of people masking 90% of the time. Masking also highlights the critical nature of suppressing super-spreader events. If nearly 100% compliance could be achieved among persons with 10 or more exposure contacts per day, then this would be sufficient for maintaining R_e_ less than one. This result highlights that policies mandating the proper use of masks at all times by all persons at sites of known super-spreader events including high risk work environments, locker rooms, weddings, social gatherings, and schools should be considered.

Slight increases in mask efficacy could also drive R_e_ to much lower levels. We believe that different types of masks should be comparatively tested with the same scientific rigor applied to clinical trials of small molecular agents and vaccines.

An important artifact of widespread masking is that while the total number of incident cases is expected to decrease dramatically, the proportion of transmissions in which at least one member of the transmission pair is masked will be higher. Therefore, anecdotal documentation of successful infections between masked individuals, or even super-spreader events in which many infected people were masked, should not be misinterpreted as failure of masking policy. The counterfactual, that masks limited the severity of these events, is likely to be true. Only longitudinal incidence data, along with shifts in level of masking in a given region, are appropriate for inferring the effectiveness of masks.

It is unlikely that antiviral therapy will prove nearly as useful as widespread masking for preventing transmissions. We previously demonstrated that antiviral therapy given during the early symptomatic phase of infection has the potential to limit duration of shedding and infection associated inflammation and is likely to be more efficacious than therapy given later during COVID-19 to hospitalized patients (*18*). Unfortunately, early symptomatic therapy would likely occur after peak viral load in a majority of cases. Our simulations suggest that even 100% penetrance of extremely potent antiviral therapies would have a negligible impact on population level spread of the virus. Therefore, while treatment as a prevention is a vital piece of HIV public health policy (*30*), it is unlikely to impact the COVID-19 pandemic.

On the other hand, treatment in the earliest pre-symptomatic phase of infection, which could only realistically occur in the setting of PEP, happens prior to peak viral load and therefore could limit secondary transmissions. However, the gains from this approach diminish with each day following exposure. In order to meaningfully impact R_e_, over 50% of exposed contacts would need to receive fully effective therapy within 3 days of an exposure. Given that no available agent yet achieves this level of efficacy and that identifying 50% of post-exposure contacts is unrealistic in most countries, it is clear that relative to masking, PEP will only have an adjunctive role in managing the pandemic. Potential areas of implementation are among high risk populations such as skilled nursing facility residents or cancer center patients and among populations where masking is difficult or impossible.

Our prior work strongly suggests the presence of a transmission dose response curve in which exposure viral load is a key determinant of transmission risk (*32, 33*). Our current analysis is built upon this assumption. We project that masks will lead to a lower exposure viral load among newly infected people, particularly if both the transmitter and exposed individual are successfully masked. Animal models of SARS-CoV-1 and MERS (*27, 28, 34*), as well as challenge studies with influenza H1N1 in humans (*35*), all demonstrate that lower exposure dose is associated with less severe disease, and human data from the Hong Kong SARS-CoV-1 outbreak in Hong Kong suggest a similar trend (*36*). While data for SARS-CoV-2 in humans is lacking, it is notable that age-adjusted hospitalization and death rates may be decreasing since more widespread utilization of masks. A mask related reduction in exposure viral load is a plausible but unproven reason for this observation.

Our work has key limitations. First, based on available data, it is impossible to know the true average efficacy of a mask worn by a transmitter or an exposed contact. Many epidemics at the state level have demonstrated a reduction in the effective reproductive number ranging from 0.2-0.5 when more widespread masking was implemented, even as physical distancing levels waxed and waned. Our model suggests that if 50-75% of people wear masks 50-75% of the time, which is roughly in accordance with state level observations of mask compliance, then a broad estimate for real world mask efficacy is ∼0.4-0.6, assuming that efficacy is equal between transmitters and exposed contact, and that masks are properly used to optimize their efficacy. If transmitter masking is more efficacious, while exposed contact masking is proportionally less efficacious, then similar results can be expected. The real-world estimate is inclusive of multiple factors including variability in mask type and masking technique. Regardless of the precise estimate, it is clear that wider implementation would yield significant reductions in spread of SARS-CoV-2 at the population level.

Second, our generalized model is not region-specific for the current pandemic. The relative impact of super-spreader events, intensity of transmission and proportion of symptomatic cases may vary from region to region based on contact network structure and age demographics. Nevertheless, the general qualitative conclusions about masking are insensitive to these differences and are likely to be generalizable across the globe.

Finally, our model does not include a standard SIR format and therefore does not capture other dynamic features that might alter the force of infection such as herd immunity or time-variant shifts in degree of physical distancing. Another missing feature that could be captured with an SIR modeling framework is the possibility of an assortative mixing pattern, in which individuals with lower adherence to masking might preferentially interact with others who have low adherence to masking (*12*). Such an effect could allow persistence of SARS-CoV-2 within this sub-population, even if masking is sufficient for containment in the rest of the population.

In conclusion, we developed a mechanistic model to demonstrate how masks reduce exposure SARS-CoV-2 viral load and transmission probability. Widespread use of even modestly effective masks is predicted to severely limit epidemic spread and represents the key available intervention along with physical distancing, to mitigate the number of infections, and perhaps the proportion of infections that are severe, while the world awaits a widely available and effective vaccine.

## Methods

### SARS-CoV-2 within-host model

We used the within-host model describing the SARS-CoV-2 infection from our previous study (*18*). This model assumes that the contact of SARS-CoV-2 (*V*) with susceptible cells (*S*) produces infected cells at rate *βVS* which then generates new virus at a per-capita rate *π*. The model also incorporates the death of infected cells mediated by (1) the innate responses (*δI*^*k*^) and (2) the acquired immune responses 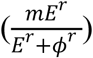 by SARS-CoV-2-specific effector cells (*E*). The magnitude of the innate immunity is dependent on the infected cell density and the exponent *k*. The nonlinearity of the acquired responses is captured by the Hill coefficient *r* that allows for rapid saturation of the killing. Finally, the parameter *ϕ* defines level of SARS-CoV-2-specific effector cells at which the killing of infected cells becomes half maximal. In the model, the rise of SARS-CoV-2-specific effector cells rise is described in a two-stage manner. The first stage defines the proliferation of the first precursor cell compartment (*M*_1_) at rate *ωIM*_1_ and differentiation into a second precursor cell compartment (*M*_2_) at a per capita rate *q*. Finally, second precursor cells differentiate into effector cells at the same per capita rate *q* and are cleared at rate *δ*_*E*_.

The model is expressed as a system of ordinary differential equations:

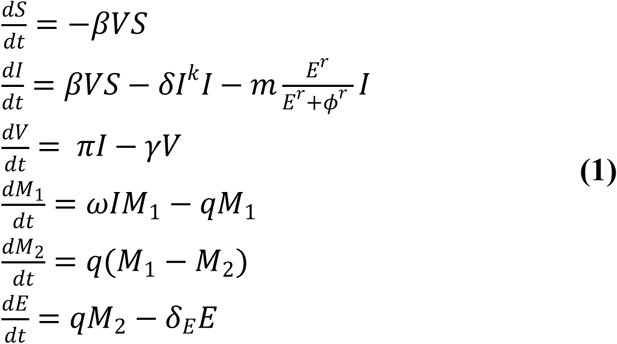

The initial conditions for the model were assumed as *S*(0) = 10^7^ cells/mL, *I*(0) = 1 cells/mL, 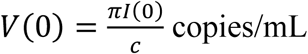 copies/mL, *M*_1_(0) = 1, *M*_2_(0) = 0 and *E*_0_ = 0. For simulations we sampled parameter values from a nonlinear mixed-effect model as described in (*19*), with the following fixed effects and standard deviation of the random effects (in parenthesis): Log_10_*β*: - 7.23 (0.2) virions^-1^ day^-1^; *δ*: 3.13 (0.02) day^-1^ cells^-k^; *k*: 0.08 (0.02); Log10(*π*): 2.59 (0.05) day^-1^; *m*: 3.21 (0.33) days^-1^cells^-1^; Log10(*ω*): −4.55 (0.01) days^-1^cells^-1^. We also assumed *r* = 10; *δ*_*E*_ = 1 day^-1^; *q* = 2.4 × 10^−5^ day^-1^ and *c* = 15 day^-1^.

### Dose-response model

We employed our previously developed dose-response model to estimate the probability of virus entering the airway given a transmitter viral load (i.e., contagiousness) and the probability of cellular infection given a transmitter viral load, (i.e., infectiousness) *P*_*t*_[*V*(*t*)] (response) based on viral loads *V*(*t*) (dose) (*19*). The relation between the response and the dose follows, 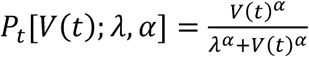, being *λ* the viral load that corresponds to 50% infectiousness and 50% contagiousness and *α* the Hill coefficient that controls the sharpness in the dose-response curve. We assumed that the viral load-dependent contagiousness (i.e., the probability that virus is passaged to the exposed person’s airway) is the same as infectiousness. We estimate the transmission risk as the product of the infectiousness and contagiousness (*19*).

### Transmission model and reproduction number

As in our previous model (*19*), we determined the total exposed contacts of a transmitter within a time step (Δ_*t*_) using a gamma distribution, i.e. 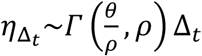, where *θ* and *ρ* represent the average daily contact rate and the dispersion parameter, respectively. The true number of exposure contacts (with viral airway exposure) was then obtained by multiplying the total exposed contacts and the contagiousness of the transmitter (i.e., 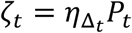). We modelled infectiousness as a Bernoulli event with mean *P*_*t*_, yielding the number of secondary infections within a time step as 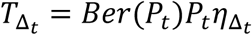. Finally, we summed up the number of secondary infections over 30 days since the time of exposure to obtain the individual reproduction number, i.e. 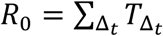. For each successful transmission, we further assumed that it takes *τ* days for the first infected cell to produce virus.

In simple steps, we followed the procedure below to estimate R_e_,

1. Simulate viral load *V*(*t*) of a simulated infected individual using **the within-host model**.
2. For a given combination of (*λ, τ, α, θ, ρ*)
  a. For each time step Δ_*t*_
    i. Compute *P*_*t*_[*V*(*t*); *λ, α*]
    ii. Draw 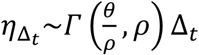
    iii. Calculate 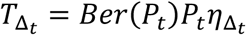
  b. Calculate 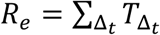
3. Repeat Steps 1 and 2 to estimate *R*_*e*_ for 3,000 infected individuals. The population level *R*_0_ can then be calculating by taking the mean of 3,000 individual *R*_*e*_ values.

### Parameter values for the transmission model

For simulations, we used the parameter set [*α, λ, τ, θ, ρ*] = [0.8, 10^7^, 0.5, 4, 40]) as they most closely reproduces empirically observed individual *R*_0_ and serial interval histograms as well as mean *R*_0_ across individuals (*R*_0_ ∈ [1.4, 2.5]) and mean serial interval across individuals (SI ∈ [4.0, 4.5]) early during the pandemic (*20-22, 24, 25*).

### Modeling mask use

To evaluate the impact of the use of mask on epidemics, we first assumed that a mask decreases the exposure viral load by a fraction (1 − *ϵ*), being *ϵ* the mask efficacy or the proportion of viruses filtered by the mask of transmitter or exposed individuals. If the transmitter is wearing mask with efficacy *ϵ*_*T*_ and the exposed person is wearing a mask with efficacy *ϵ*_*E*_, then the combined mask efficacy *ϵ*_*C*_, is given by 1 − (1 − *ϵ*_*T*_)(1 − *ϵ*_*E*_). Infectiousness or contagiousness reduction by the use of mask can be computed as:

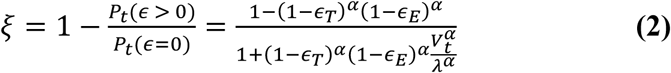

Similarly, the transmission risk reduction using mask can be computed as:

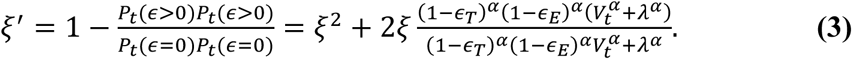

Finally, we modeled the *compliance* of an individual wearing mask in the population as a Bernoulli event with mean *σ* and the *adherence* of wearing it at time step Δ_*t*_ as a Bernoulli event with mean *ϑ*. Compliance is defined as whether the person ever wears a mask. Adherence is the percentage of time that a mask wearer wears a mask.

### Simulating secondary transmissions with mask use

For a specific scenario with selected *σ* and *ϑ*, we followed the procedure below to estimate the population level *R*_0_:

1. Simulate *V*(*t*) for a transmitter using the within-host model in **eq. 1**.
2. Simulate transmitter mask compliance using *Ber*(*p* = *σ*).
3. Discretize the time-space of 30 days over time steps Δ_*t*_. For each time step,
  a. If the transmitter is *compliant* in using a mask:
    i. Simulate transmitter mask *adherence* at time step Δ_*t*_ using *Ber*(*p* = *ϑ*)
    ii. If transmitter is wearing a mask at time step Δ_*t*_
      1. Draw 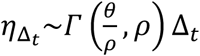.
      2. Determine masking adherence among exposed contacts at time step Δ_*t*_ using *k*∼*Ber*(*p* = *σϑ*). If *k* = 1, then there is 100% adherence among exposed contacts and if *k* = 0, then there is 0% adherence among exposed contacts.
      3. Determine the number of exposed contact wearing a mask (i.e., 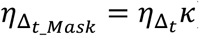) and the number of exposed contact not wearing a mask (i.e., 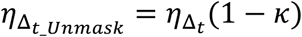.
        A. Compute 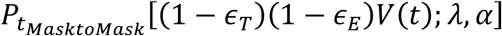
        B. Calculate 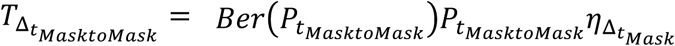
        C. Compute 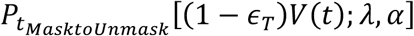
        D. Calculate 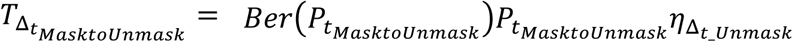
    iii. If the transmitter is not adhering with the use of a mask at time step Δ_*t*_, which is determined at step (i).
      1. Draw 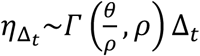.
      2. Determine masking adherence among exposed contacts at time step Δ_*t*_ using *k*∼*Ber*(*p* = *σϑ*).
      3. Determine the number of exposed contact wearing a mask (i.e., 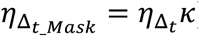) and the number of exposed contact not wearing a mask (i.e., 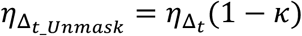
        A. Compute 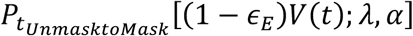
        B. Calculate 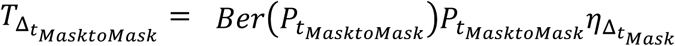
        C. Compute 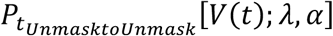
        D. Calculate 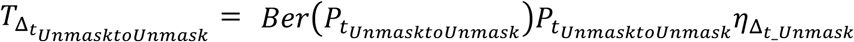
  b. If the transmitter is not compliant in wearing a mask
    1. Draw 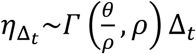.
    2. Determine masking adherence among exposed contacts at time step Δ_*t*_ using *k*∼*Ber*(*p* = *σϑ*).
    3. Determine the number of exposed contact wearing a mask (i.e., 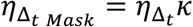) and the number of exposed contact not wearing a mask (i.e., 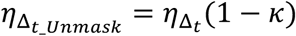
      A. Compute 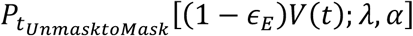
      B. Calculate 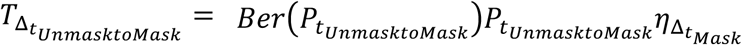
      C. Compute 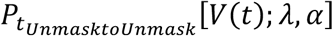
      D. Calculate 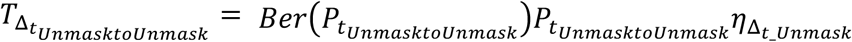
  c. Calculate 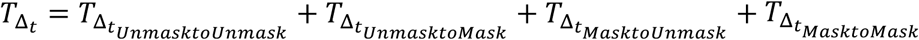.
4. Calculate 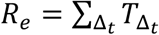.
5. Repeat Steps **1** to **4** to estimate *Q*_5_ for 3,000 infected individuals (transmitters). R_e_ can then be calculating by taking the mean of 3,000 individual *Q*_5_ values.

### Modeling antiviral treatment

We simulate the antiviral treatment by assuming that the antiviral treatment reduces the viral production (*π*) by (1 − *ϵ*_*treat*_), where *ϵ*_*treat*_ is the efficacy of treatment. Here, *ϵ*_*treat*_ = 0 and *ϵ*_*treat*_ = 1 represent the case of completely ineffective and 100% effective treatment, respectively. For the transmission simulations we model *coverage* to treatment with as a Bernoulli event with mean *ψ*.

In the presence of treatment with mean coverage *ψ* and efficacy *ϵ*_*treat*_, we follow the procedure below to estimate the population level *R*_*e*_:

1. Determine adherence to treatment using *Ber*(*p* = *ψ*)
2. Determine the time of start of antiviral treatment (*T*_*treat*_) for treatment in symptomatic phase and pre-symptomatic phase by randomly drawing a number from Uniform distributions (*U*(0.5 + *I*_*I*_, 5*_I*_*I*_)) and (*U*(0.5,5)), respectively, where *I*_*I*_ is the incubation period of the infected individual.
3. Simulate viral load *V*(*t*) of a simulated infected individual using the within-host model in **eq. 1** with *ϵ*_*treat*_ = 0 for *t* ≤ *T*_*treat*_ and 0 < *ϵ*_*treat*_ ≤ 1 for *t* > *T*_*treat*_.
4. For a given combination of (*λ, τ, α, θ, ρ*)
  a. For each time step Δ_*t*_
    i. Compute *P*_*t*_[*V*(*t*); *λ, α*]
    ii. Draw 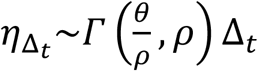
    iii. Calculate 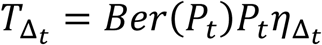
  b. Calculate 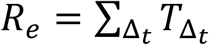
5. Repeat Steps 1 and 4 to estimate *R*_*e*_ for 3,000 infected individuals. The population level *R*_*e*_ can then be calculating by taking the mean of 3,000 individual *R*_*e*_ values.

## Data Availability

All modeled data and code will be available upon request.

---

**Supplementary figure 1.**
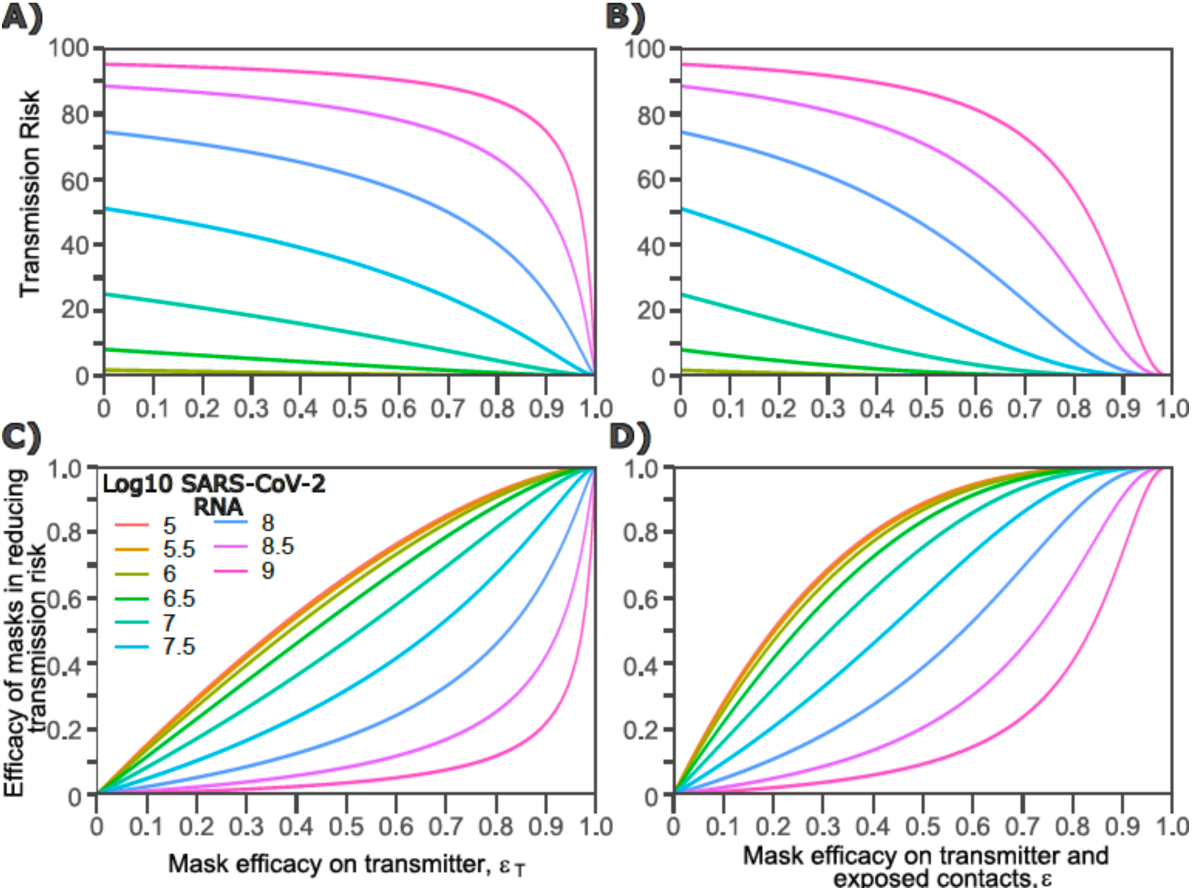
Impact of masking of the transmitter or exposed contact alone or both members of the transmission pair, on transmission risk given an exposure viral load. Each panel is based on simulations of 1000 transmission pairs. **a-b**. Absolute reduction in transmission risk. **c-d**. Relative reduction in transmission risk. **a**,**c**. Transmitter is masked only **b, d**. Both transmitter and exposed contact are masked

**Supplementary figure 2.**
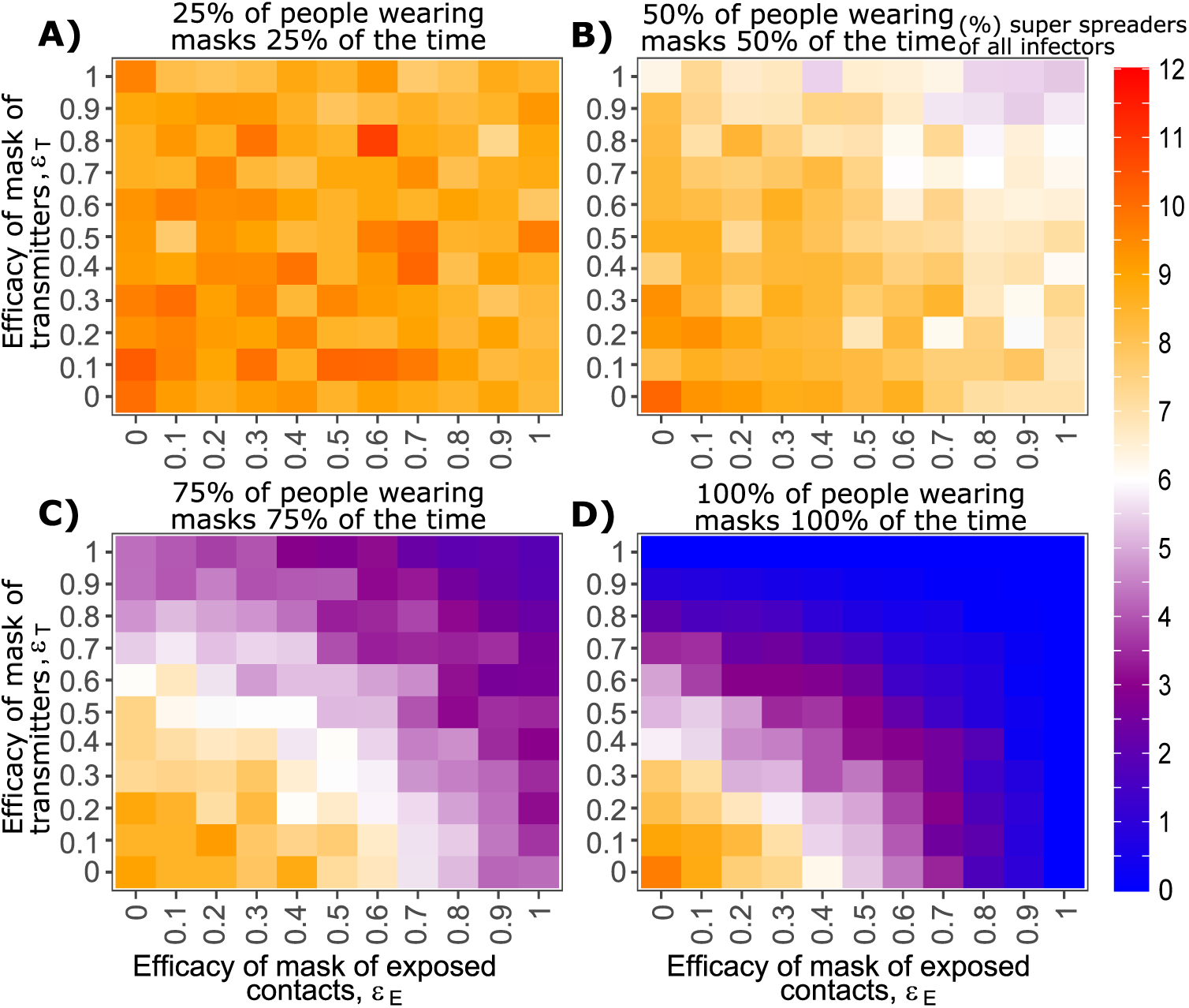
Effect of mask utilization and efficacy on percent of infectors who supers-spread to others. Each heat map is based on simulations of 3000 transmitters with varying daily exposure contacts. **a**. Low mask utilization, **b**. Moderate mask utilization, **c**. High mask utilization, **d**. Extremely high mask utilization.

**Supplementary figure 3.**
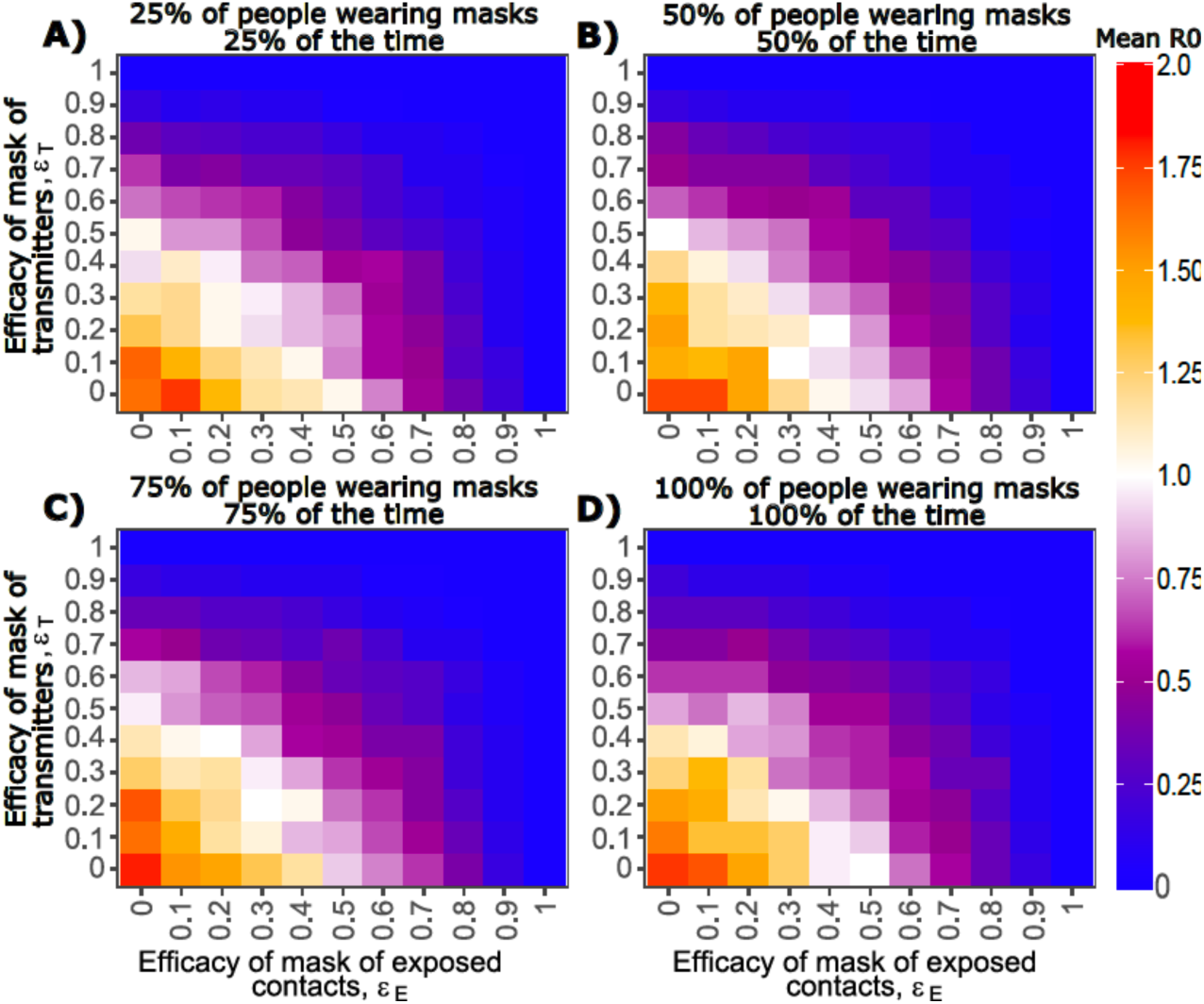
Effect of mask utilization and efficacy on mean SARS-CoV-2 R0 assuming all persons with >10 exposure contacts per day are masked. Each heat map is based on simulations of 3000 transmitters with varying daily exposure contacts. **a**. Low mask utilization, **b**. Moderate mask utilization, **c**. High mask utilization, **d**. Extremely high mask utilization.

**Supplementary figure 4.**
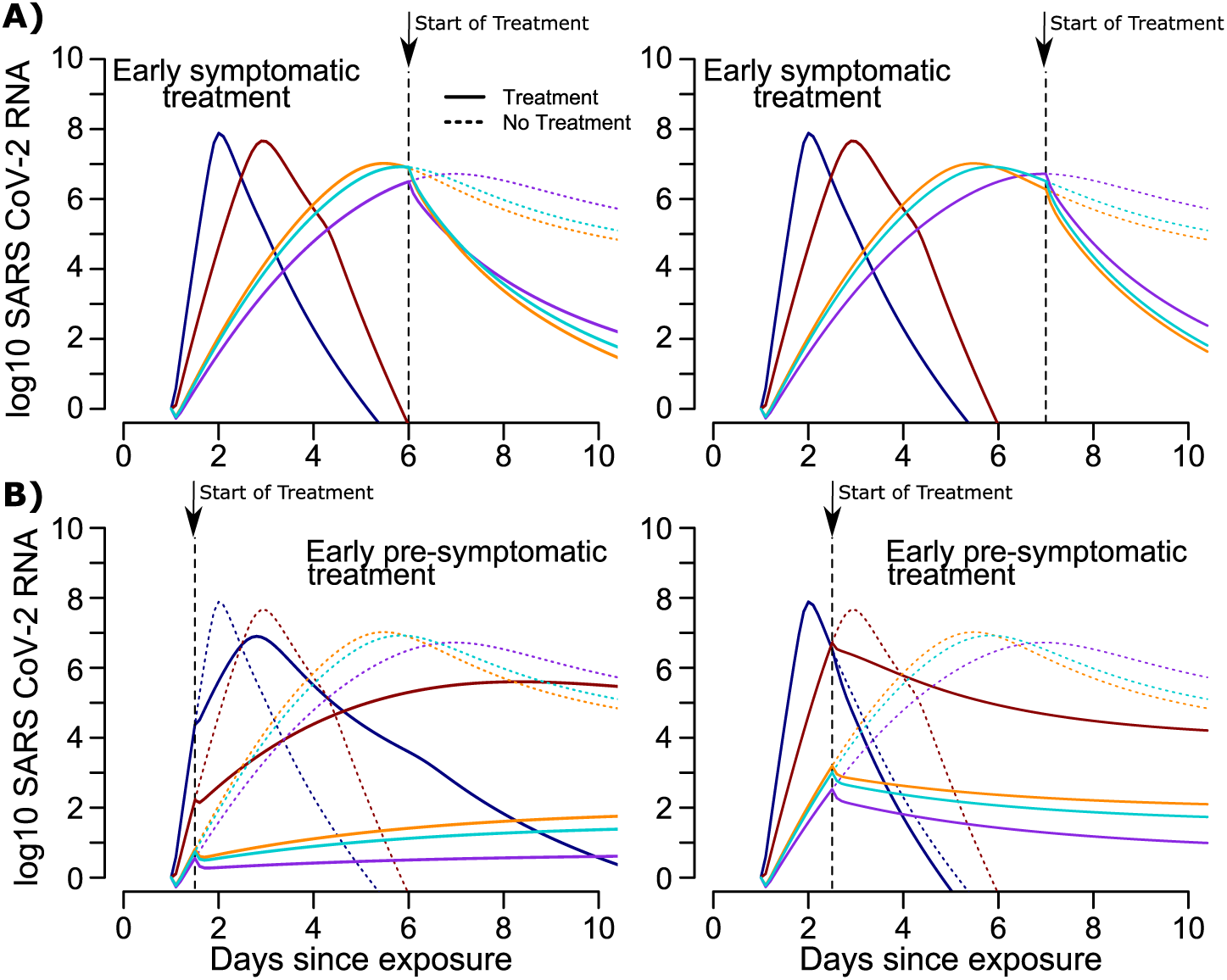
Viral load simulations on antiviral therapy. **a-b.**Solid lines trajectories are assumed with therapy, whereas dashed lines are counterfactuals with no therapy. **a**. Simulated early symptomatic therapy. **b**. Simulated PEP.

